# Non-Traditional Lipid Ratios Predict Cardiovascular-Kidney-Metabolic Syndrome: Insights from Machine Learning Model Using NHANES Data

**DOI:** 10.1101/2025.09.26.25336774

**Authors:** Nan Wang, Jie Zhang, YuYing Wang, ZhaoXia Zhang, Ya Shi, Lu Zhang, Ying Xing, QianQian Lv

## Abstract

**Background:** Cardiovascular-kidney-metabolic (CKM) syndrome is a newly defined multisystem disease continuum characterized by the coexistence of metabolic dysfunction, cardiovascular disease, and chronic kidney disease (CKD). This study aimed to explore the relationship between the non-traditional lipid ratios and CKM syndrome.

**Methods:** A cross-sectional analysis was performed using data from the 2007-2018 National Health and Nutrition Examination Survey (NHANES). CKM stages (0-4) were classified according to the 2023 American Heart Association (AHA) criteria. Non-traditional lipid ratios, including triglyceride to high-density lipoprotein cholesterol (TG/HDL-C), total cholesterol to HDL-C (TC/HDL-C), low-density lipoprotein cholesterol to HDL-C (LDL-C/HDL-C), and non-HDL-C, were assessed. Nonlinear associations were explored using restricted cubic spline models. Variable selection was conducted using the least absolute shrinkage and selection operator (LASSO) regression, and four machine learning models were developed. SHapley Additive exPlanations (SHAP) and partial dependence plots (PDPs) were applied for interpretability.

**Results:** The significant associations were observed between non-traditional lipid ratios and CKM risk. Nonlinear relationships were observed for LDL, TG, TC, and HDL. The random forest (RF) algorithm demonstrated superior performance. SHAP analysis identified body mass index (BMI) as the most influential predictor, followed by TG, TG/HDL-C, and glucose. Elevated TG/HDL-C, TC/HDL-C, and non-HDL-C levels were positively associated with increased CKM risk, while LDL-C/HDL-C showed inverse associations. PDPs indicated synergistic interactions among TG/HDL-C, TC/HDL-C, and non-HDL-C. Multivariate logistic regression confirmed TG/HDL-C as an independent predictor (OR = 3.11; 95% CI: 2.49-3.88; *p* < 0.001).

**Conclusion:** TG/HDL-C and non-HDL-C are strong predictors of CKM syndrome. Integration of these markers into risk models may facilitate earlier detection and guide individualized prevention strategies.

## Introduction

Cardiovascular-kidney-metabolic (CKM) syndrome has recently been recognized as a systemic clinical entity that reflects complex interactions among obesity, diabetes, chronic kidney disease (CKD), and cardiovascular disease (CVD)^1^. Unlike traditional single-organ frameworks, CKM highlights multi-organ crosstalk, where metabolic disturbances accelerate cardiovascular and renal dysfunction, while organ impairments further intensify metabolic derangements. CKM has been linked to renal failure^2^, metabolic dysfunction-associated steatotic liver disease (MASLD)^3,4^, and increased risks of certain cancers^5,6^. Epidemiological evidence suggests that about 5% of U.S. adults already exhibit coexisting cardiovascular, renal, and metabolic conditions, with prevalence rising steadily, underscoring its growing public health burden^7^.

Global Burden of Disease (GBD) analyses have further emphasized the significance of CKM syndrome. Disability-adjusted life years (DALYs) attributable to seven core components of CKM syndrome have increased steadily between 1990 and 2021, driven largely by population growth and aging. Projected trends indicate that stroke of DALYs will increase by 55.9%, and the burden of atrial fibrillation and flutter by 105.7% from 2022 to 2046^8^. These projections underscore the need for early detection, risk stratification, and preventive strategies, as CKM syndrome is poised to remain a major contributor to global morbidity.

Dysregulated lipid metabolism has emerged as a pivotal mechanism in CKM syndrome. Conventional lipid metrics, including total cholesterol (TC), low-density lipoprotein cholesterol (LDL-C), and high-density lipoprotein cholesterol (HDL-C), are routinely employed for cardiovascular risk assessment but often lack sufficient predictive accuracy in the context of multi-morbidity. In contrast, non-traditional lipid parameters, derived from conventional lipid measures, have demonstrated enhanced predictive value for complex cardiometabolic disorders. Parameters such as the Atherogenic Index of Plasma (AIP)^9^, non-HDL cholesterol (non-HDL-C)^10^, and the Cholesterol Retention Index (CRI-I and CRI-II)^11^ integrate multiple lipid fractions, reflecting both atherogenic potential and systemic metabolic disturbances.

Epidemiological evidence confirms that non-traditional lipid indices are strongly associated with predicting cardiometabolic multimorbidity (CMM) and its components^12–14^. Elevated AIP correlates with increased cardiovascular and type 2 diabetes risk, suggesting utility as an early metabolic marker. Non-HDL-C and CRI indices demonstrate stronger associations with metabolic syndrome than LDL-C. Mechanistic studies indicate that these parameters are linked to inflammation, oxidative stress, and insulin resistance (IR)^15–17^. Adipose tissue contributes further to pathogenesis: as an endocrine organ, it secretes adipokines, cytokines, and free fatty acids that impair insulin signaling, promote chronic inflammation, and accelerate metabolic deterioration^18,19^. These processes affect cardiovascular, renal, and hepatic function, reflecting the multi-organ progression of CKM syndrome.

Despite these insights, research remains limited by a focus on individual disease components rather than CKM syndrome as an integrated entity. The combined predictive value and potential synergistic effects of non-traditional lipid indices are underexplored, constraining their clinical translation for early identification and risk stratification. Given their ability to detect metabolic abnormalities earlier than conventional measures, systematic investigation is warranted.

This study aims to evaluate the associations between non-traditional lipid parameters and CKM syndrome and to assess their predictive utility using machine learning. Integrating multiple lipid indices permits consideration of cross-organ interactions and the identification of high-risk individuals at early stages of disease progression. Findings are expected to elucidate shared mechanisms of organ damage, inform novel intervention targets, and provide evidence for the simultaneous prevention of cardiovascular, renal, and metabolic disorders, addressing a growing global health burden.

## Methods

All data are publicly available and can be accessed at the NHANES website (https://wwwn.cdc.gov/nchs/nhanes/Default.aspx). Relevant R code is available upon reasonable request to the corresponding author.

### Data sources and study population

A cross-sectional analysis was conducted using data from the NHANES from 2007 to 2018. NHANES employed a multistage, stratified probability sampling design to generate a nationally representative sample of the non-institutionalized U.S. population. Health-related data were collected through standardized interviews, physical examinations, and laboratory testing^20^. The inclusion criteria were as follows: 1) adults aged ≥ 18 years were included; 2) Exclusion criteria consisted of pregnancy, lactation; 3) missing critical variables, or inability to undergo cardiovascular risk evaluation. A total of 4,689 participants were eligible and randomly assigned to training (n = 3,282) and validation (n = 1,407) sets at a 7:3 ratio (Figure 1).

**Figure 1.**
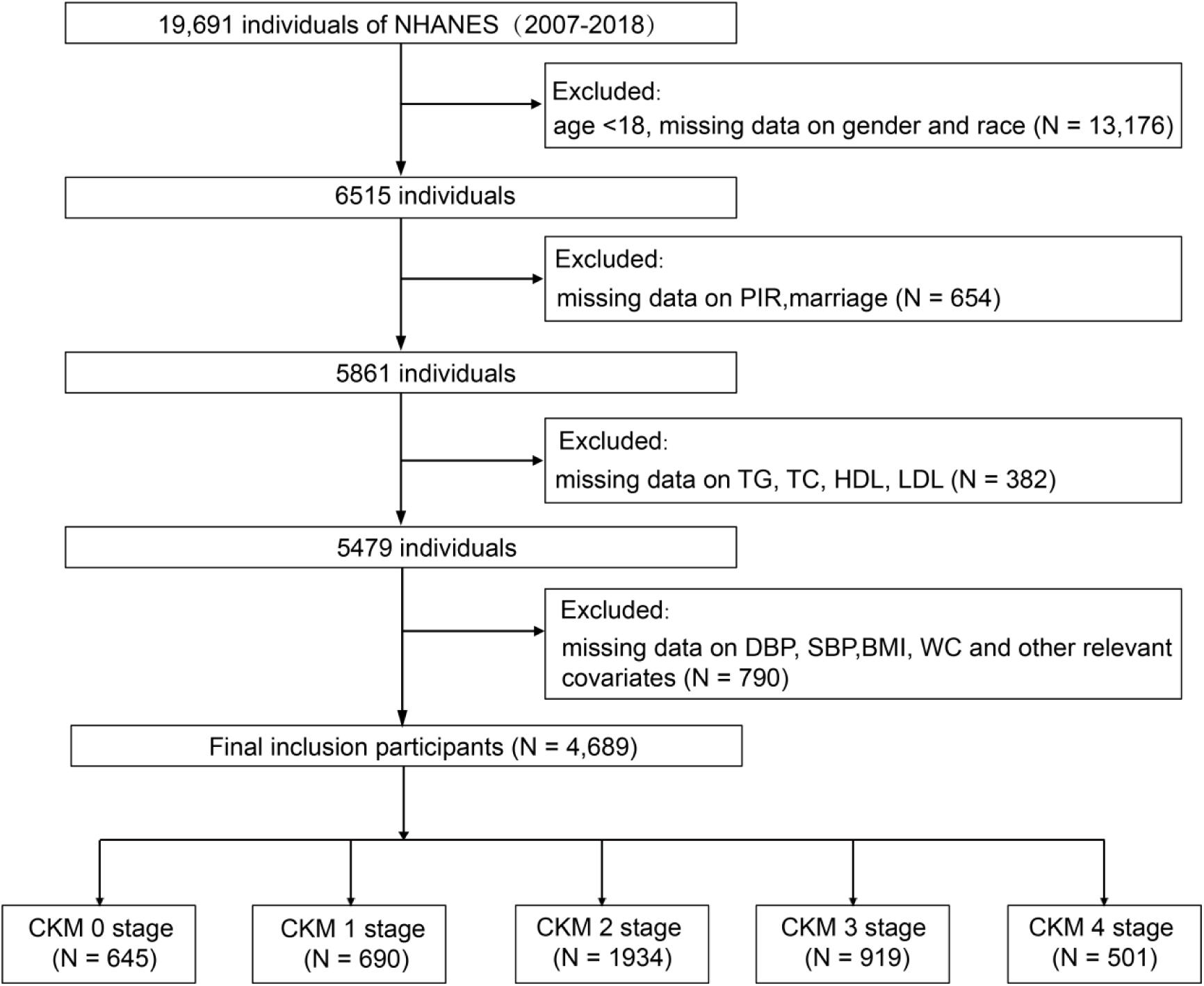
Study participant selection process from NHANES 2007-2018.

### Definition of exposure

Exposure variables included four lipid-related biomarkers, including triglyceride to high-density lipoprotein cholesterol ratio (TG/HDL-C), total cholesterol to HDL-C ratio (TC/HDL-C), low-density lipoprotein cholesterol to HDL-C ratio (LDL-C/HDL-C), and non-HDL-C. The HDL-C was measured using direct immunoassay or precipitation methods. The TC and HDL-C were measured enzymatically using the Hitachi 704 Analyzer (Boehringer Mannheim Diagnostics, Indianapolis, IN, USA). LDL-C was calculated using the Friedewald equation^21^. Non-HDL-C was derived by subtracting HDL-C from TC (Supplementary Table S1). All laboratory measurements were obtained from the NHANES laboratory database and standardized by the U.S. Centers for Disease Control and Prevention.

### Definition of cardiovascular-kidney-metabolic syndrome

The primary outcome was CKM syndrome, defined by the 2023 American Heart Association staging criteria. CKM stages were determined based on three dimensions, including metabolic dysfunction, CVD, and CKD^22^. Stage 0 indicated no CKM risk, stage 1 included individuals with obesity or impaired glucose metabolism, stage 2 involved additional metabolic disorders or moderate-to-high CKD risk, stage 3 reflected subclinical CVD, and stage 4 indicated established clinical CVD (Supplementary Table S2). Ten-year CVD risk was estimated using the AHA PREVENT equations, applicable to adults aged 30–79 years. High CVD risk was defined as a predicted 10-year risk ≥20%. CKD staging was determined based on estimated glomerular filtration rate (eGFR), calculated using the race-neutral CKD-EPI 2021 creatinine equation, and urinary albumin-to-creatinine ratio (ACR).

### Covariates

Covariates included demographic variables (age, sex, race, education level, marital status, and poverty income ratio (PIR)), lifestyle behaviors (smoking, alcohol use), chronic conditions (hypertension, diabetes, heart disease, angina, myocardial infarction, stroke), and laboratory biomarkers (body mass index (BMI), albumin, and creatinine) (Supplementary Table S3).

### Statistical analysis

Baseline characteristics were summarized for the total sample, training, and validation sets. Continuous variables were reported as weighted means with standard errors (SE), and categorical variables as weighted percentages. Between-group differences were evaluated using weighted t-tests^23^. Restricted cubic spline (RCS) models were applied to assess nonlinear associations between lipid ratios and CKM risk, with *p*-overall and *p*-nonlinear used to test significance^24^.

To address high-dimensional input variables, the least absolute shrinkage and selection operator (LASSO) regression was used for feature selection^25^. The optimal penalty parameter was determined using 10-fold cross-validation to minimize mean squared error. Generalized variance inflation factors were calculated to evaluate multicollinearity, values below 10 were considered acceptable.

Four supervised machine learning methods were developed using the training set, including random forest (RF), support vector machine (SVM), decision tree (DT), and multilayer perceptron (MLP)^26^. Data preprocessing included one-hot encoding for categorical variables and standardization for continuous variables. Hyperparameters were optimized using grid search and cross-validation. Model performance was assessed using area under the curve (AUC), accuracy, sensitivity, specificity, and F_1_ score. Confusion matrices were generated to visually compare classification results, and the model with the highest overall performance was selected.

To interpret model output, SHapley Additive exPlanations (SHAP) were used to quantify each variable’s contribution^27^. Variable importance rankings and SHAP dependency plots were created to assess directional influence. Partial dependence plots (PDPs) were applied to examine synergistic effects and variable interactions^28^. To quantify the independent association between the key lipid biomarker and CKM syndrome, multivariable logistic regression models were employed, where participants were dichotomized into two groups: Stage 0 (no CKM risk) versus Stages 1-4 (presence of CKM risk)^29^. Specifically, Model 1 (unadjusted) used univariate logistic regression to assess the crude association of key lipid biomarkers with CKM syndrome, while Model 2 (adjusted) used multivariable logistic regression, adjusting for sex, age, education level, and hypertension status. Results for both models were expressed as odds ratios (ORs) with corresponding 95% confidence intervals (95% CI). This logistic regression analysis was performed separately on both the training set and the validation set.

All analyses were conducted in R (version 4.4.1) and Python (version 3.12.3), with visualizations produced using “ggplot2” and “matplotlib”. A two-sided *p*-value <0.05 was considered statistically significant.

## Results

### Baseline characteristics of participants

A total of 4,689 participants were included and stratified by CKM syndrome stage. Among the overall population, 50.8% were female, and 59.8% were aged ≥45 years (Table 1). Non-Hispanic White individuals comprised 51.1%. Statistically significant differences (*p* < 0.05) were observed across CKM stages in sociodemographic factors (age, sex, race, marital status, education, PIR), lifestyle behaviors (smoking, alcohol use), comorbidities (hypertension, diabetes, heart disease, angina, stroke), and biochemical indicators (serum creatinine, HDL-C, TC). Although the proportion of females was balanced across most stages, it declined to 41.3% in Stage 4. The proportion of individuals aged ≥45 years increased to 94.8% in Stage 4. Non-Hispanic White participants were most prevalent in Stage 4 (64.5%), and this stage also had the highest proportion of married individuals (55.5%). Higher education levels (College graduate or above) decreased with CKM progression, dropping from 36% in Stage 0 to 18.2% in Stage 4. The proportion of non-drinkers increased from 20.2% to 28.9% in Stage 4. Serum albumin declined from 4.38 g/dL to 4.14 g/dL, creatinine increased from 70.89 to 95.08 mg/dL, BMI from 22.03 to 30.14 kg/m², and glucose from 93.02 to 119.50 mg/dL (Table 1). In the training set, all variables differed significantly by CKM stage; in the validation set, all but depression showed significance (Supplementary Table S4-S5).

**Table 1.**
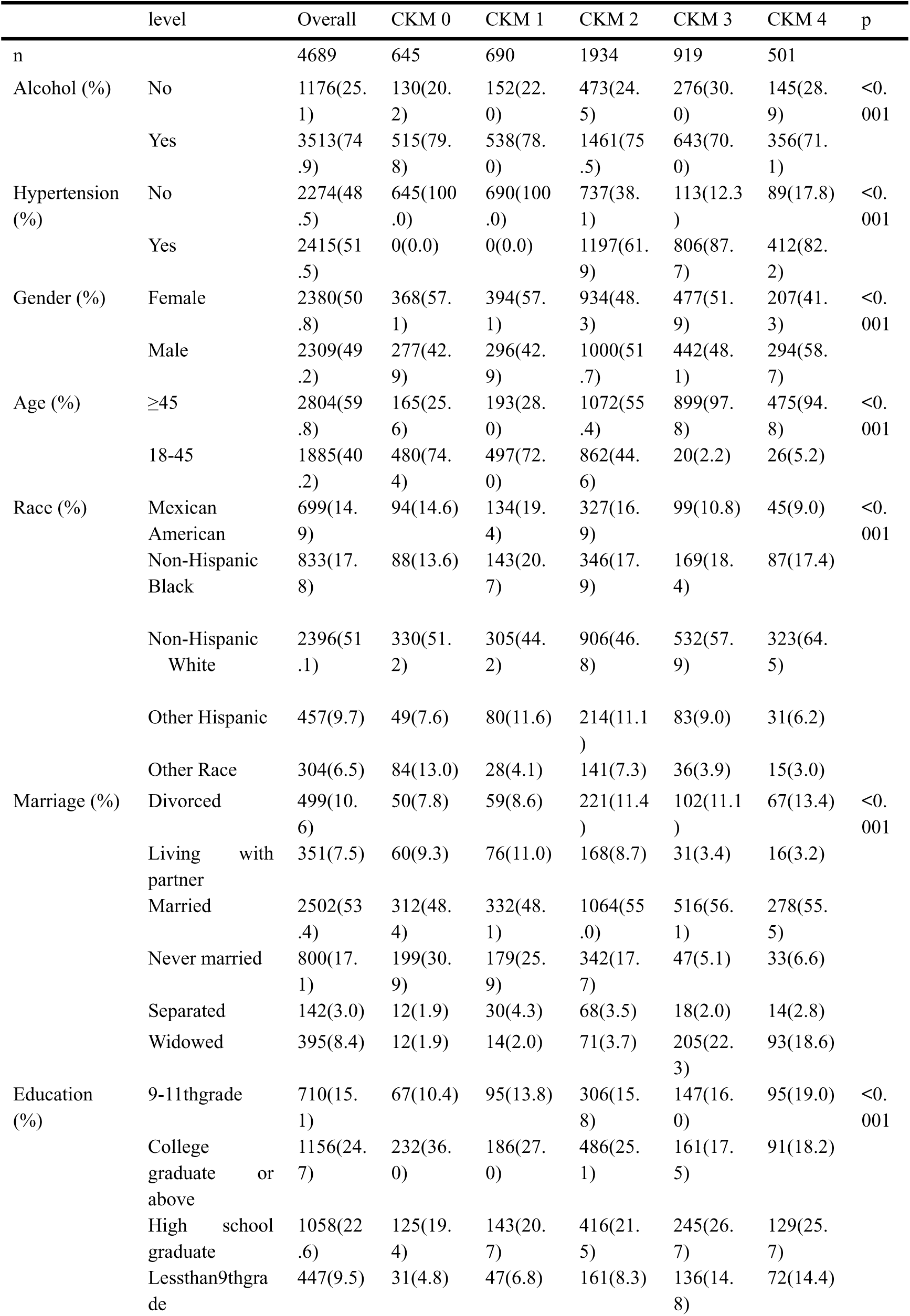
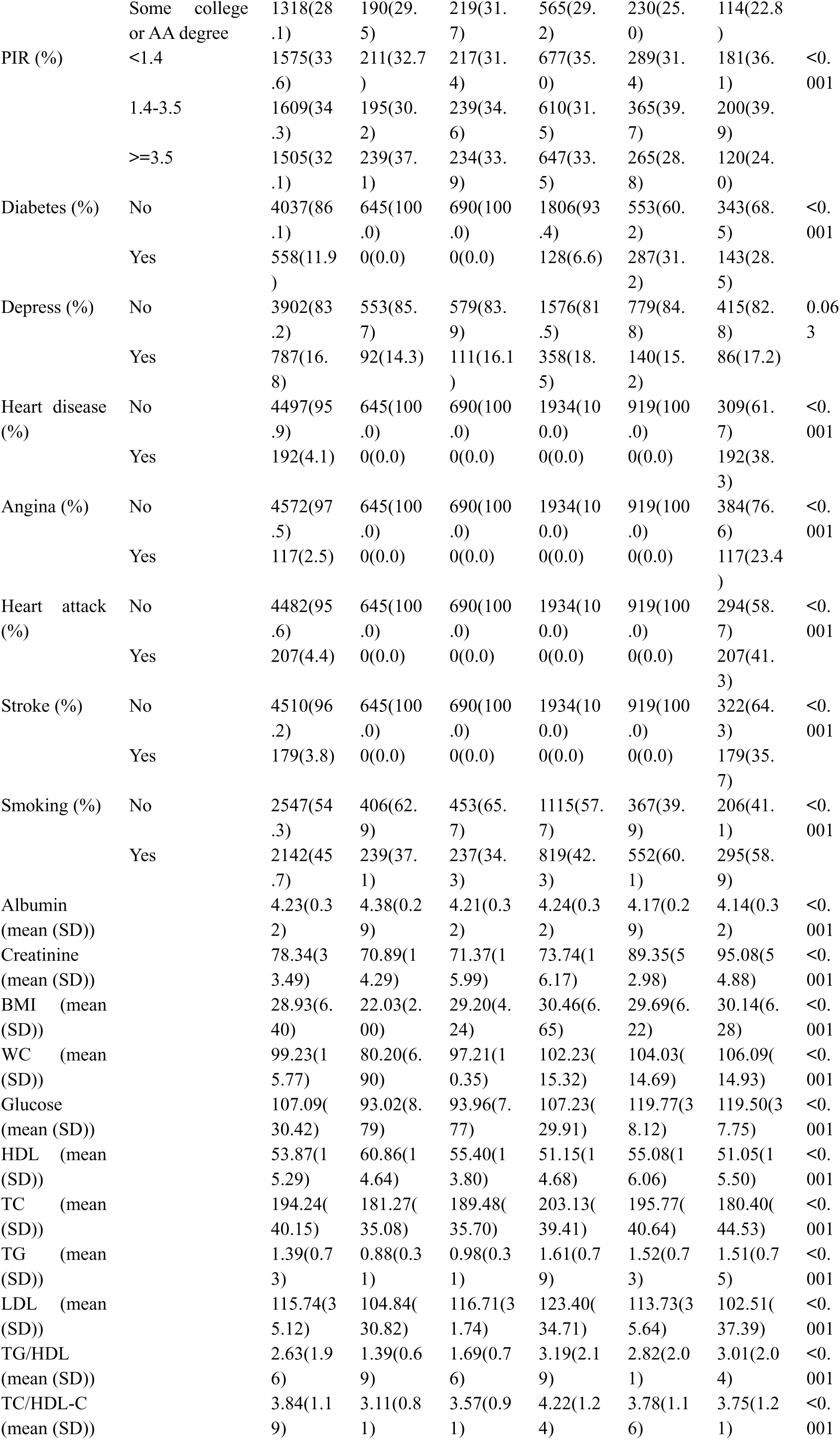
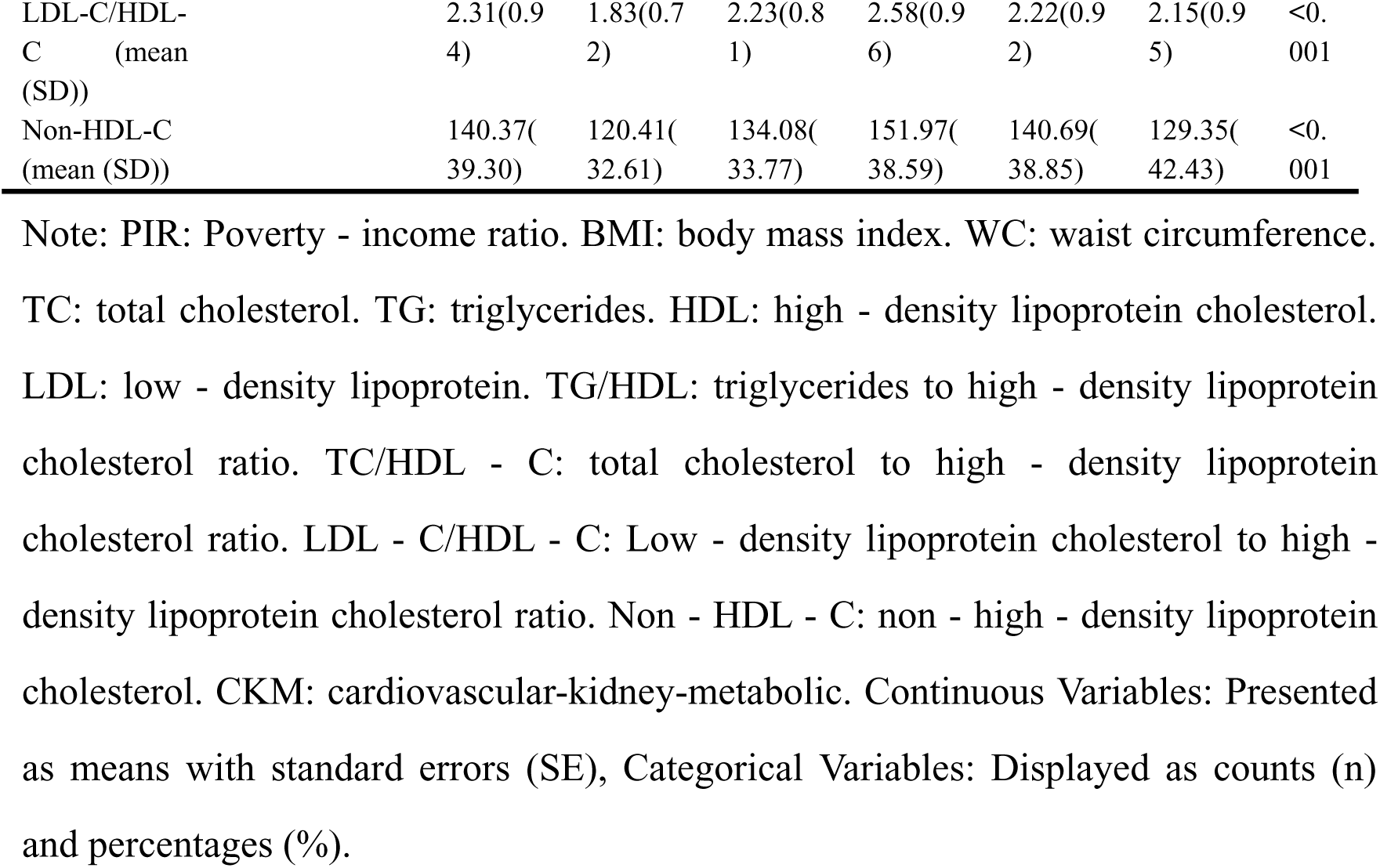
Baseline characteristics of the total study population according to different stages of CKM syndrome.

### Nonlinear associations between lipid markers and CKM risk

The RCS identified significant nonlinear relationships between CKM risk and lipid markers, including low-density lipoprotein (LDL) (*p*-overall < 0.01; *p*-nonlinear < 0.01), TG (*p*-overall < 0.01; *p*-nonlinear < 0.01), and TC (*p*-overall < 0.01; *p*-nonlinear = 0.0208). Increased levels of glucose, TG/HDL-C, TC/HDL-C, LDL-C/HDL-C, and non-HDL-C were positively associated with CKM risk (*p*-overall < 0.05), whereas higher high-density lipoprotein cholesterol (HDL) was inversely associated (*p*-overall < 0.01; *p*-nonlinear < 0.01) (Figure 2).

**Figure 2.**
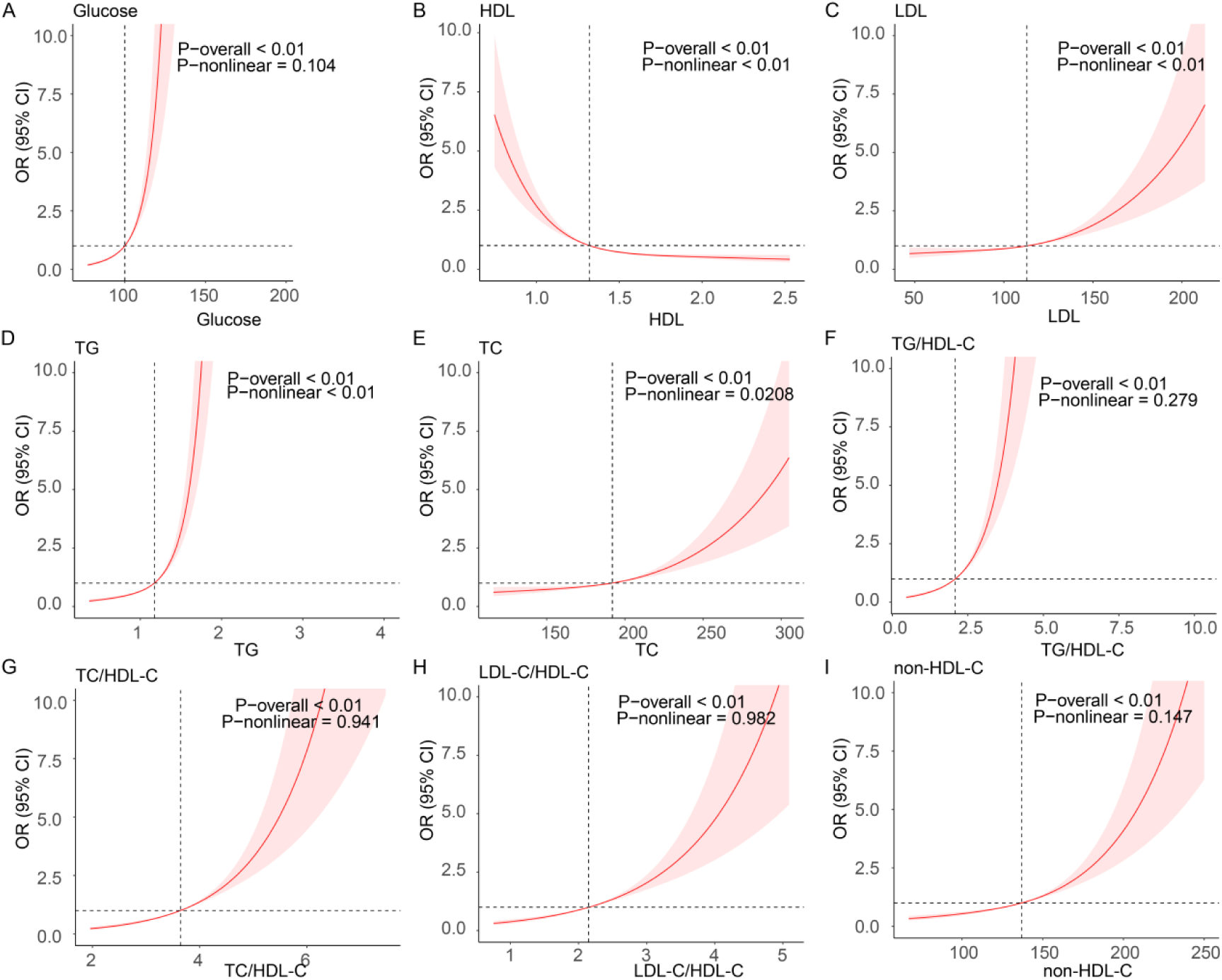
RCS curves of significant exposure variables associated with CKM syndrome. (A) Glucose. (B) HDL. (C) LDL. (D) TG. (E) TC. (F) TG/HDL-C. (G) TC/HDL-C. (H) LDL-C/HDL-C. (I) non-HDL-C.

### Feature selection and machine learning model performance

The association between cross-validation error and the log-transformed penalty parameter (λ) in LASSO regression analyses was used to identify exposures that are more relevant to CKM syndrome, and the 24 exposures included in the LASSO regressions had the smallest error in the model at λ = 3×10^-5^, with all variables retained, suggesting that all variables are potentially relevant (Figure 3).

**Figure 3.**
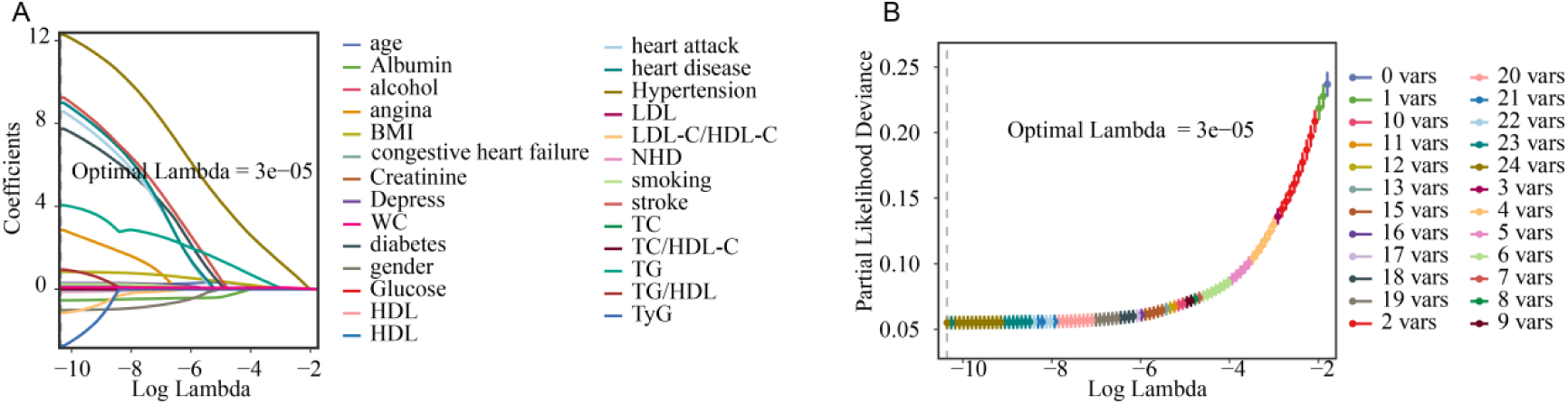
LASSO screening of characterization variables. (A) LASSO coefficient paths. Horizontal axis: log(lambda); vertical axis: variable coefficients. As lambda increases, coefficients shrink toward zero. Variables with zero coefficients are excluded at the optimal lambda. (B) LASSO cross-validation. Horizontal axis: log(lambda); vertical axis: model error. Optimal lambda minimizes model error.

The RF model demonstrated superior performance, with an AUC of 0.97, accuracy of 93.106%, sensitivity of 0.953, and specificity of 0.784. In comparison, the DT (AUC = 0.84), SVM (AUC = 0.94), and MLP (AUC = 0.93) models in the training set showed lower specificity. The confusion matrix for the RF model showed 1,165 true positives, 57 false positives, 145 true negatives, and 40 false negatives, confirming its suitability as the final predictive model (Figure 4, Supplementary Table S6).

**Figure 4.**
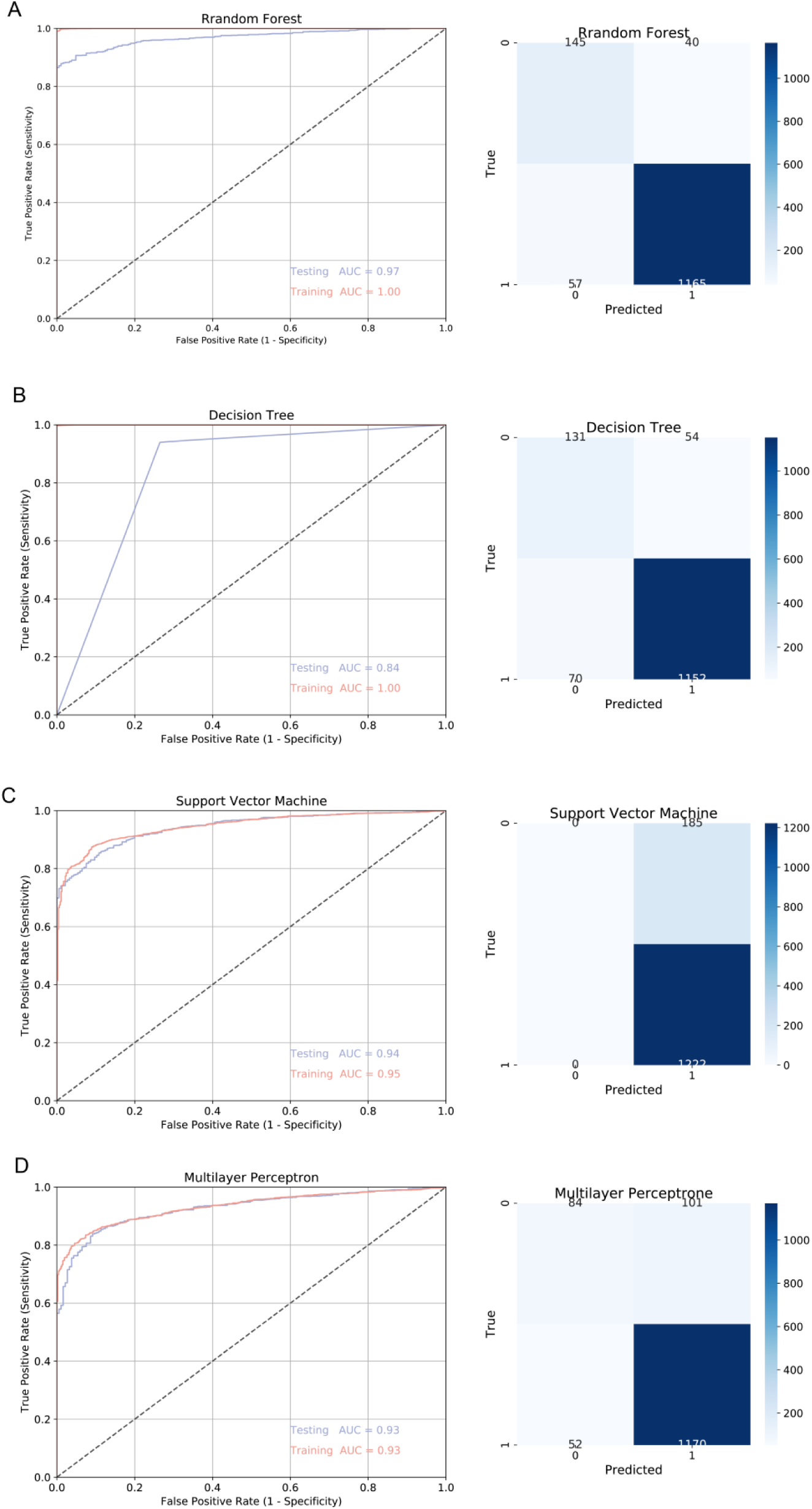
ROC curves of the test and train sets and mixing matrices. (A) RF. (B) DT. (C) SVM. (D) MLP.

### Variable importance in the RF model

Variable importance analysis revealed that BMI had the highest predictive contribution (importance = 0.454), followed by TG (0.100), TG/HDL-C (0.097), and glucose (0.095). In contrast, TC (0.036) and LDL (0.035) demonstrated lower contributions (Figure 5A).

**Figure 5.**
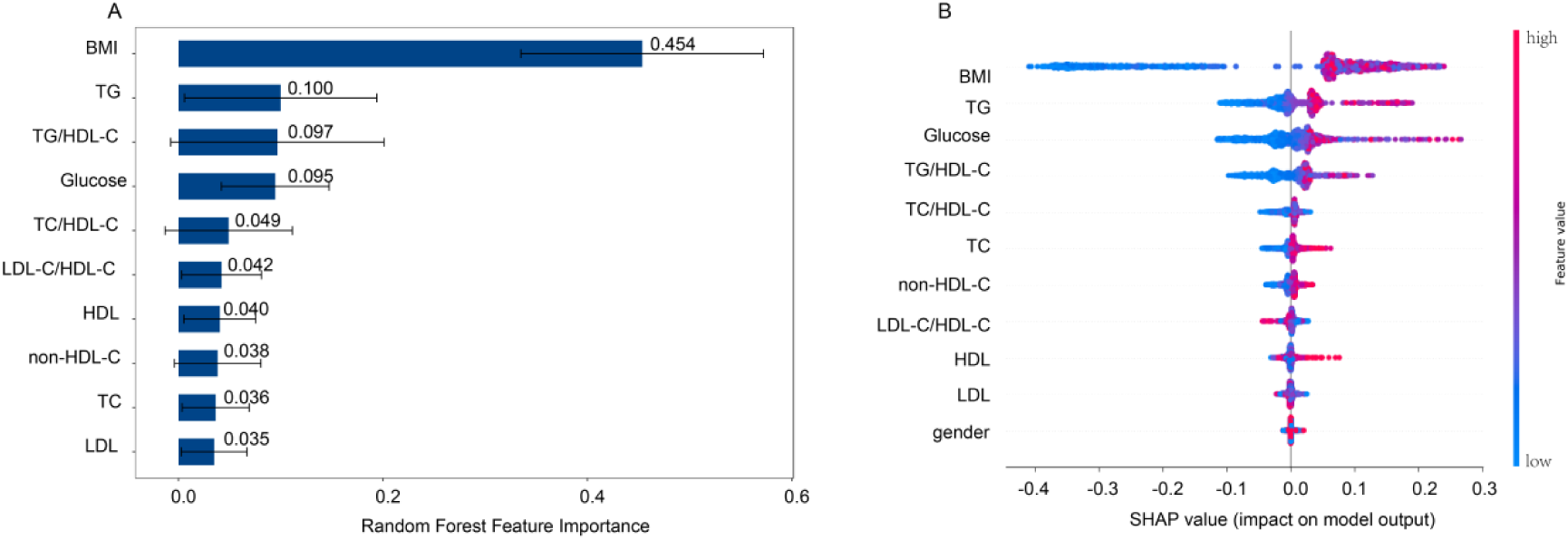
RF model. (A) Characteristic importance of RF models. (B) The effect of each feature in the predictive model on the CKM syndrome was explained by the SHAP value in the RF models.

### Relationships between the lipid markers and CKM syndrome

PDPs showed that TG/HDL-C and non-HDL-C levels were associated with increased CKM risk. A nonlinear pattern was observed for TC/HDL-C, which initially increased with risk and then declined. An inverse association was noted for LDL-C/HDL-C (Supplementary Figure S1).

### Synergistic effects of lipid markers and CKM syndrome

The synergistic effects revealed that TG/HDL-C and TC/HDL-C synergistically elevated predicted CKM risk, with TG/HDL-C playing a dominant role. Further synergistic effects between TC/HDL-C and LDL-C/HDL-C, as well as between LDL-C/HDL-C and non-HDL-C, which significantly affect CKM syndrome risk estimation. Lower LDL-C/HDL-C and higher non-HDL-C levels were consistently linked to elevated CKM syndrome risk (Supplementary Figure S2).

### Model decision of SHAP

The SHAP analysis indicated that elevated TG/HDL-C, TC/HDL-C, and non-HDL-C levels were associated with higher SHAP values, suggesting a greater CKM risk. The SHAP value for TG/HDL-C plateaued at higher values. TC/HDL-C showed stable contributions at mid-range levels but fluctuated at extremes. Non-HDL-C exhibited a rising trend that eventually stabilized, while LDL-C/HDL-C showed decreasing SHAP values with increasing levels (Supplementary Figure S3).

SHAP importance rankings further confirmed BMI as the strongest predictor. The TG and TG/HDL-C were positively associated with risk, while glucose and TC/HDL-C demonstrated a negative association (Figure 5B).

### Association between TG/HDL-C and CKM syndrome

Logistic regression analysis conducted in both the training set and validation set (n = 645 for Stage 0; n = 4,044 for Stages 1-4) confirmed TG/HDL-C as a robust independent predictor of CKM syndrome. In unadjusted models (model 1), significant associations were observed in the training (OR = 3.11; 95% CI: 2.49-3.88; *p* < 0.01) and validation sets (OR = 3.10; 95% CI: 2.39-4.01; *p* < 0.01). After adjustment for sex, age, education level, and hypertension (model 2), the association remained statistically significant in the testing (OR = 3.39; 95% CI: 2.52-4.54; *p* < 0.01), confirming its independent predictive value (Figure 6).

**Figure 6.**
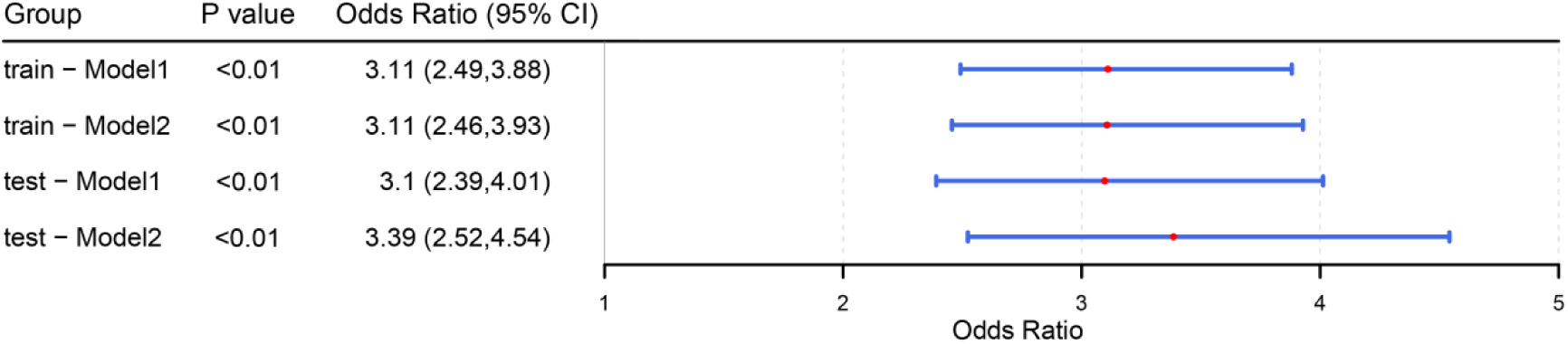
Forest plots comparing model 1 and model 2 between TG/HDL-C and CKM syndrome. Note: contains only the TGH univariate model; Model 2: based on model 1, 4 indicators of sex, age, education, and Hypertension were added to the training set.

## Discussion

A comprehensive evaluation of non-traditional lipid parameters revealed their strong association with CKM syndrome, offering critical insights into both pathophysiology and potential early identification of this multi-organ disorder. Elevated TG/HDL-C and non-HDL-C were identified as key contributors to CKM risk, with TG/HDL-C emerged as a strong predictor of CKM risk when combined with TC/HDL-C. Predictive models incorporating these indices achieved superior performance, highlighting the non-linear interactions among lipid fractions that traditional linear analyses fail to capture. SHAP analysis further confirmed TG/HDL-C as a robust independent predictor, exhibiting stabilized influence at higher values and aligning with its pathophysiological relevance in cardiometabolic dysregulation. Logistic regression analysis reinforced its independent predictive capacity across both training and validation cohorts, with adjusted ORs remaining significant, emphasizing its potential utility for early risk stratification.

Dyslipidemia remains a principal driver of CKM syndrome, with triglycerides and LDL-C recognized as key determinants of CVD risk and prognosis, whereas HDL-C exerts protective effects through cholesterol efflux, endothelial stabilization, and antioxidant activity^30–32^. Conventional lipid metrics, though routinely employed in clinical practice, insufficiently capture the complex metabolic interactions underpinning multi-organ pathophysiology in CKM. Non-traditional lipid parameters integrate multiple lipid fractions, providing a more comprehensive reflection of systemic metabolic disturbance and atherogenic potential^33–36^. These observations align with prior cohort studies demonstrating that elevated TG/HDL-C, non-HDL-C, and cholesterol retention indices are associated with increased incidence of CVD, type 2 diabetes (T2D), and CKD. Longitudinal data from the NAGALA cohort and the Korean National Health Examination Survey further demonstrated that non-traditional lipid indices independently predict incident diabetes and, when combined with abdominal obesity, substantially increase the cumulative risk of CMM^33^.

Mechanistic evidence supports the role of “pan-lipotoxicity” in CKM pathogenesis, extending beyond traditional triglyceride-centered lipotoxicity. Dysregulated adipose tissue function, ectopic lipid deposition, and abnormal secretion of adipokines and bioactive metabolites collectively contribute to multi-organ injury^37,38^. Elevated TG levels activate cholesteryl ester transfer protein (CETP), reducing HDL-C while increasing LDL-C, thereby generating small dense LDL particles with enhanced endothelial permeability and pro-inflammatory potential^39,40^. Dyslipidemia contributes to both atherosclerosis and thrombogenesis, exacerbates insulin resistance, and promotes metabolic syndrome progression, amplifying cardiovascular and renal injury^41,42^. Renal pathophysiology in CKM is tightly linked to lipid-mediated damage, excessive cholesterol and phospholipid reabsorption in proximal tubules triggers tubulointerstitial inflammation, foam cell formation, and fibrotic remodeling, while intracellular cholesterol accumulation impairs podocyte and tubular cell function, perpetuating CKD progression^43–46^. Reciprocal interactions between declining renal function and lipid dysregulation create a feed-forward loop that exacerbates multi-organ deterioration^47^. These findings provide a mechanistic rationale for the observed superior predictive value of TG/HDL-C in identifying individuals at risk for CKM syndrome.

Recent studies emphasize the interplay between lipid abnormalities and subclinical cardiovascular dysfunction. Elevated TG/HDL-C correlates with endothelial impairment, reduced vascular compliance, and chronic low-grade inflammation, contributing to early atherogenesis and left ventricular remodeling^35,36^. Concurrent derangements in non-HDL-C and TC/HDL-C reflect cumulative atherogenic burden and residual risk not captured by LDL-C^10,11^. Multi-parametric lipid evaluation thus enables earlier identification of cardiometabolic derangements and high-risk individuals prior to overt CKM manifestation. The additive and synergistic effects of lipid indices further underscore the importance of integrated metrics rather than reliance on single measures for risk assessment.

From a clinical perspective, incorporating non-traditional lipid parameters into risk assessment frameworks represents a paradigm shift. Tree-based machine learning algorithms leveraged the complex interactions among lipid indices to enhance predictive accuracy. SHAP and partial dependence analyses elucidated individual contributions as well as synergistic interactions, providing mechanistic insight into their predictive utility^12^. Such approaches highlight the limitations of linear modeling in capturing the multi-dimensional and non-linear nature of CKM, supporting the adoption of advanced computational methods for comprehensive risk stratification. Moreover, machine learning outputs offer interpretable metrics that could facilitate personalized intervention strategies, such as identifying individuals most likely to benefit from targeted lipid-lowering therapies or lifestyle modification.

Policy and public health implications are substantial. With the global prevalence of metabolic syndrome, CKD, and CVD continuing to rise, early identification of high-risk populations using integrative lipid metrics could inform preventative strategies, including structured lifestyle interventions, pharmacotherapy addressing dyslipidemia and insulin resistance, and individualized monitoring programs. Population-level screening programs incorporating non-traditional lipid indices may facilitate the detection of subclinical CKM, enabling timely intervention to prevent progression to end-organ damage^34,48^. Such strategies align with contemporary guidelines emphasizing multi-dimensional risk assessment and personalized management in metabolic and cardiovascular disorders.

This study provides several novel contributions. First, it represents one of the largest analyses examining non-traditional lipid parameters in relation to CKM syndrome, establishing TG/HDL-C as a particularly robust predictor. Second, it employs machine learning methodologies to model complex non-linear relationships among lipid fractions and disease outcomes, providing a framework for future integrative risk prediction tools. Third, by combining clinical, laboratory, and demographic variables, the study offers a holistic perspective on early cardiometabolic and renal risk that could be leveraged in both clinical and public health contexts. Furthermore, mechanistic interpretations grounded in pan-lipotoxicity and lipid-mediated endothelial and renal injury support the biological plausibility of the observed associations, bridging epidemiologic findings with underlying pathophysiology.

Several limitations warrant consideration. First, CVD risk was estimated using the Framingham 10-year risk score, which may not fully capture all subclinical pathologies or account for emerging risk factors. Second, cardiovascular outcomes were self-reported, potentially introducing measurement bias and underreporting. Third, the study population was limited to U.S. adults, which may reduce the generalizability of findings to other ethnicities or geographic regions. Fourth, as an observational analysis, causality cannot be inferred, and residual confounding by unmeasured factors, including dietary patterns, genetic predisposition, and inflammatory biomarkers, cannot be entirely excluded. Fifth, molecular biomarkers, advanced imaging, and metabolomic data were unavailable, limiting mechanistic exploration. Future research should leverage prospective multi-ethnic cohorts, integrate multi-omics, inflammatory and endothelial biomarkers, and utilize imaging to validate non-traditional lipid parameters in CKM risk prediction and to elucidate pathways linking dyslipidemia, insulin resistance, and multi-organ injury.

Emerging areas for further investigation include the interaction of non-traditional lipid parameters with gut microbiota, systemic inflammation, and oxidative stress pathways, which may modulate cardiometabolic risk in CKM populations. In addition, evaluating the longitudinal trajectories of lipid indices in relation to CKM progression could inform dynamic risk prediction models. The potential benefits of interventions targeting TG/HDL-C and non-HDL-C for multi-organ protection remain largely unexplored, warranting randomized clinical trials and mechanistic studies. Integrating lifestyle, pharmacologic, and precision medicine approaches based on these predictive indices may optimize early prevention strategies, mitigate end-organ damage, and reduce long-term healthcare burden.

## Conclusion

In summary, BMI emerges as the key predictive factor for CKM, accounting for 0.454 of the predictive power. Additionally, the TG/HDL-C (0.097) and glucose (0.095) further refine risk stratification. Analysis indicates a positive correlation between TG/HDL-C and non-HDL-C with the occurrence of CKM, while the LDL-C/HDL-C shows a significant negative association. SHAP values and PDPs reveal that high TG/HDL-C and high TC/HDL-C synergistically promote CKM. Therefore, future CKM screening systems could consider incorporating the TG/HDL-C ratio, particularly for individuals with a BMI greater than 25 kg/m^2^. However, prospective studies are needed to validate causality and explore precise intervention strategies.

## Data Availability

All data are publicly available and can be accessed at the NHANES website (https://wwwn.cdc.gov/nchs/nhanes/Default.aspx).

## Acknowledgments

The authors thank the National Center for Health Statistics of the Centers for Disease Control and Prevention for sharing the NHANES data.

